# Novel Large Empirical Study of Deep Transfer Learning for COVID-19 Classification Based on CT and X-Ray Images

**DOI:** 10.1101/2024.08.08.24311683

**Authors:** Mansour Almutaani, Turki Turki, Y-h. Taguchi

**Affiliations:** Department of Computer Science, King Abdulaziz University, Jeddah 21589, Saudi Arabia; Department of Physics, Chuo University, Tokyo 112-8551, Japan

**Keywords:** COVID-19, medical imaging, classification, deep transfer learning, model generation, advanced AI concepts

## Abstract

The early and highly accurate prediction of COVID-19 based on medical images can speed up the diagnostic process and thereby mitigate disease spread; therefore, developing AI-based models is an inevitable endeavor. The presented work, to our knowledge, is the first to expand the model space and identify a better performing model among 10000 constructed deep transfer learning (DTL) models as follows. First, we downloaded and processed 4481 CT and X-ray images pertaining to COVID-19 and non-COVID-19 patients, obtained from the Kaggle repository. Second, we provide processed images as inputs to four pre-trained deep learning models (ConvNeXt, EfficientNetV2, DenseNet121, and ResNet34) on more than a million images from the ImageNet database, in which we froze the convolutional and pooling layers pertaining to the feature extraction part while unfreezing and training the densely connected classifier with the Adam optimizer. Third, we generate and take a majority vote of two, three, and four combinations from the four DTL models, resulting in 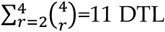 models. Then, we combine the 11 DTL models, followed by consecutively generating and taking the majority vote of 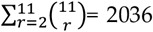 DTL models. Finally, we select 7953 DTL models from 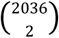. Experimental results from the whole datasets using five-fold cross-validation demonstrate that the best generated DTL model, named HC, achieving the best AUC of 0.909 when applied to the CT dataset, while ConvNeXt yielded a higher marginal AUC of 0.933 compared to 0.93 for HX when considering the X-ray dataset. These promising results set the foundation for promoting the large generation of models (LGM) in AI.

## 1. Introduction

Coronavirus disease 2019 (COVID-19) patients experience different symptoms, such as a loss of smell and/or taste, fever, headache, and shortness of breath, to name a few [1]. Human-to-human transmission can happen via (1) sneeze droplets carrying SAR-CoV-2; (2) direct contact with an infected individual; and (3) touching a virus-containing surface followed by touching the eyes or mouth [2]. To lower the risk of COVID-19 transmission, the Centers for Disease Control and Prevention suggested prevention strategies including wearing masks, social distancing, and washing hands with soap for at least 20 seconds (see Figure 1) [3, 4]. The early detection of COVID-19 and thereby isolating the infected individuals is considered the most effective prevention strategy [5, 6]. Hence, researchers have proposed various computational methods based on deep learning to detect COVID-19 using various image sources. Ozturk et al. [7] proposed a deep learning approach (DarkCovidNet) composed of interleaved convolutional and max pooling layers for feature extraction and a densely connected classifier for binary and multiclass COVID-19 classification tasks. DarkCovidNet and nine other deep learning (DL) models were applied to 635 (and 1125) X-ray test images from GitHub pertaining to binary (and multiclass) COVID-19 classification. DarkCovidNet had the highest accuracy of 0.9808 (and 0.8702) for binary (and multiclass) COVID-19 classification.

**Figure 1.**
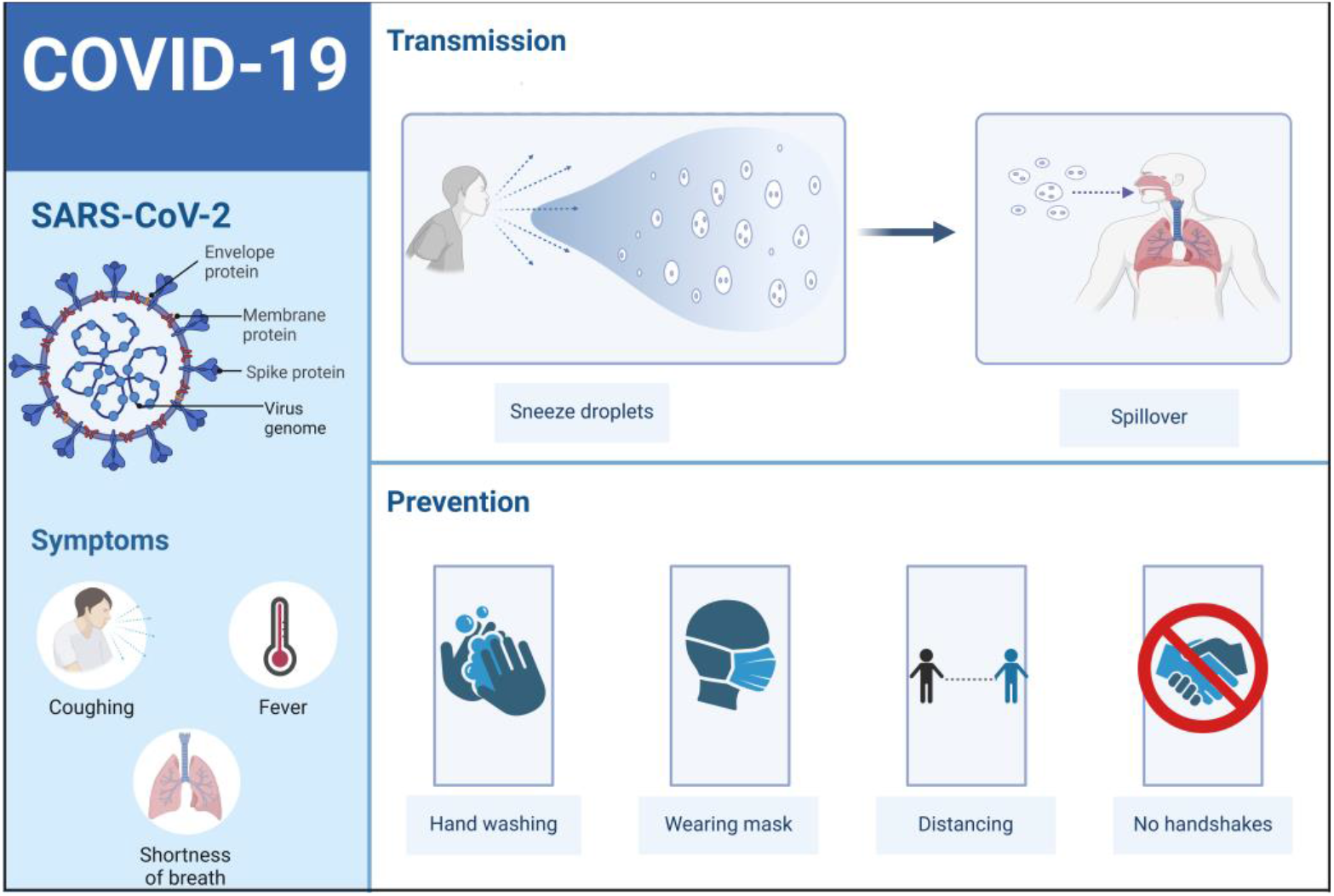
COVID-19 disease symptoms as well as transmission and prevention methods for the SARS-CoV-2 virus. Figure created with BioRender.com.

To address the COVID-19 classification task using X-ray images, Apostolopoulos et al. [8] employed five pre-trained DL models, including MobileNetV2, VGG19, Inception, Xception, and InceptionResNetV2, applied to two testing datasets obtained from GitHub and Kaggle. The first testing dataset had 1428 X-ray images for the binary class task pertaining to COVID-19 and non-COVID-19 images, while the multiclass task was to discriminate between COVID-19, pneumonia, and normal images. VGG19 achieved the highest accuracy of 0.9875 and 0.9348 for the binary and multiclass classification tasks, respectively. For the second testing dataset of 1442 test images, MobileNetv2 generated the highest accuracy of 0.9678 and 0.9472 for the binary and multiclass classification tasks, respectively. Ismael et al. [9] aimed to discriminate between normal and COVID-19 X-ray images as follows. They employed five pre-trained DL models, namely VGG16, VGG19, ResNet18, ResNet50, and ResNet101, for feature extraction, in which each was coupled with SVM to train and perform prediction. The data were downloaded COVID-19 X-ray images from GitHub, Kaggle, and Radiology Assistant. The results demonstrated that applying ResNet50 to 95 test images for feature extraction followed by SVM for classification resulted in the highest accuracy of 0.947.

Ravi et al. [10] proposed a deep and machine learning approach to discriminate between COVID-19 and non-COVID-19 X-ray and CT images. The proposed approach consisted of pre-trained EfficientNet for feature extraction followed by logistic regression as a machine learning (ML) model for classification. Compared to 26 DL models, EfficientNet + ML when applied to 2417 CT images yielded the highest accuracy of 0.9946. Moreover, EfficientNet + ML generated the highest accuracy of 0.9948 when applied to 2864 COVID-19 X-ray images. Asif et al. [11] proposed a shallow CNN trained on COVID-19 X-ray images from Kaggle. The proposed shallow CNN was compared against five pre-trained DL models (i.e., NasNet, MobileNet, Xception, InceptionV3, and DenseNet201), in which all were applied to 636 COVID-19 X-ray test images. The results demonstrated that the proposed shallow CNN achieved the highest accuracy of 0.9968. Gupta et al. [12] proposed a deep learning model (DLM) for the task of COVID-19 classification based on COVID-19 CT images from Kaggle. The proposed DLM is composed of a series of convolutional and max pooling layers for feature extraction, followed by a densely connected classifier to discriminate between COVID-10 and non-COVID-19 CT images. Compared to two pretrained models (DarkNet19 and MobileNetV2), the results demonstrated that the proposed DLM when applied to 2481 images yielded the highest accuracy of 0.9891.

Ayalew et al. [13] proposed an approach based on a CNN for feature extraction coupled with SVM for COVID-19 classification. They downloaded COVID-19 X-ray images from GitHub and then applied the CNN + SVM to 1200 X-ray images pertaining to COVID-19 and normal cases. The results demonstrated that the proposed approach achieved the highest accuracy of 0.991. Constantinou et al. [14] employed five pre-trained DL models, including InceptionV3, DenseNet169, ResNet101, DenseNet121, and ResNet50, for the multiclass COVID-19 classification task. They downloaded COVID-19 X-ray images from Kaggle. Results on 6788 test images demonstrated that ResNet101 achieved the highest accuracy of 0.96 when compared to the other DL models. In Table 1, we provide a comparison of our proposed work against existing works for COVID-19 classification.

**Table 1.**
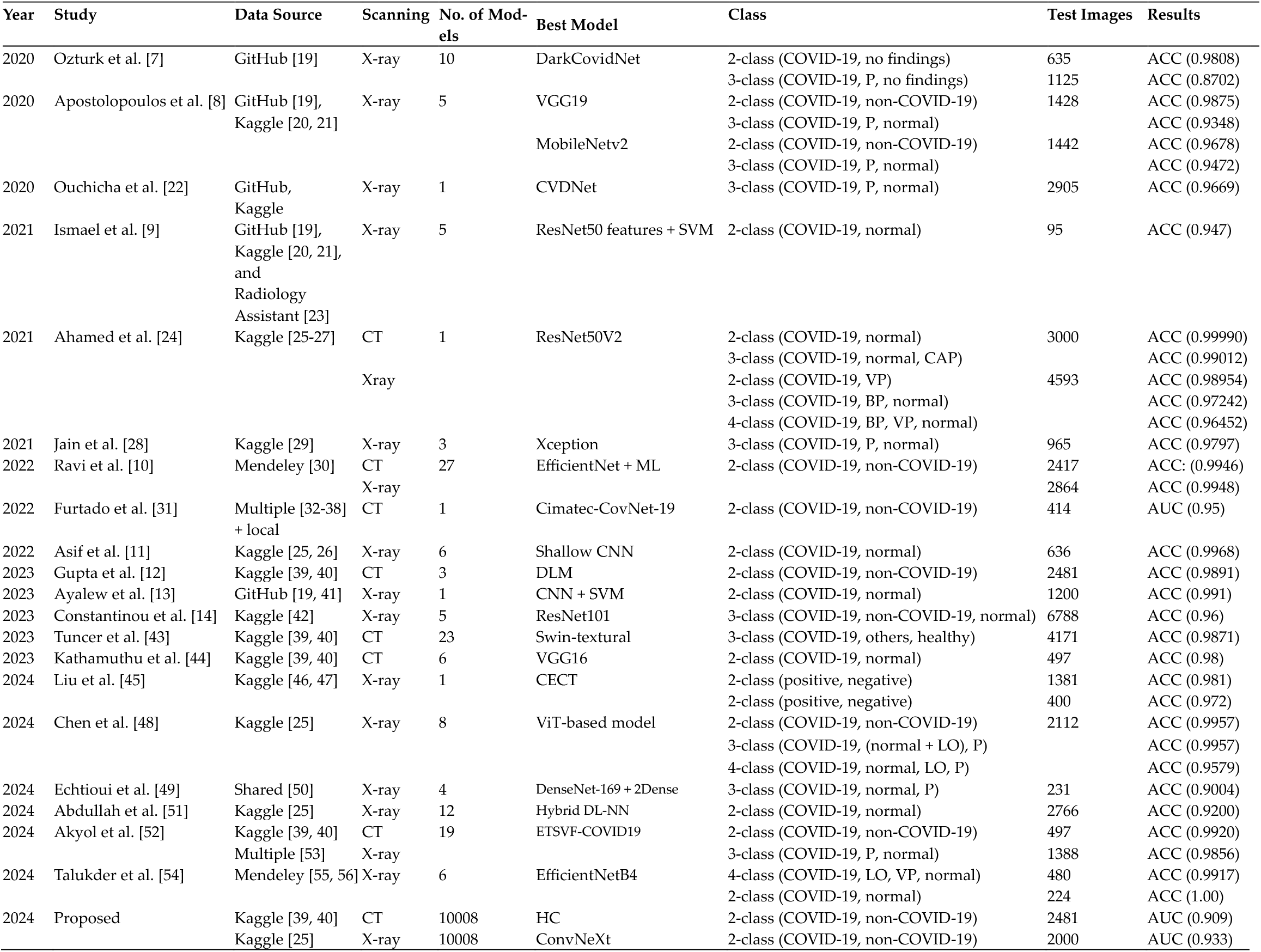
Comparison of our proposed work against other studies. P stands for pneumonia. VP stands for viral pneumonia. BP stands for bacterial pneumonia. CAP stands for community-acquired pneumonia. LO stands for lung opacity. CT is computed tomography. TEM is transmission electron microscopy. ACC is accuracy. AUC is area under the ROC curve.

These existing AI-driven computational methods with which to detect COVID-19 based on medical images are far from perfect. Therefore, automatically expanding the model space is needed to identify a better performing model improving over the performance of existing methods. Hence, we propose a computational framework for the large generation of models. The novelty in our study can be mainly ascribed to the following summarized contributions:

1. Unlike existing AI-based studies for detecting COVID-19, to our knowledge, this is the first study to investigate the large generation of AI models in which we expand the model space, constructing thousands of prediction models for the task of COVID-19 classification based on CT and X-ray images.
2. Our framework works as follows: We employed four randomly selected pre-trained DL models, including ConvNeXt, EfficientNetV2, DenseNet121, and ResNet34. Then, we downloaded 2481 and 2000 COVID-19 images from Kaggle pertaining to CT and X-ray scanning methods, respectively. For each pre-trained model, we froze the feature extraction part composed of convolutional and pooling layers while training from scratch the densely connected layer with the Adam optimizer for the binary classification, resulting in four deep transfer learning (DTL) models. We took a majority vote of two, three, and four combinations from the four DTL models, resulting in a total of 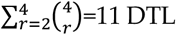 models. Next, we took 2, 3, …, and 11 combinations from the 11 DTL models, resulting in 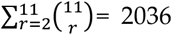 DTL models. Finally, we chose 7953 DTL models from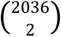.
3. According to our large empirical study of 10008 models attributed to 11 + 2036 + 7953 generated DTL models plus 8 DTL models, the generated model, named HC, which corresponds to the composition of DTL models obtained \\ the highest AUC of 0.909 when COVID-19 CT images were used. For the COVID-19 X-ray images, ConvNeXt and the generated model, HX, obtained an AUC of 0.933 and 0.930, respectively.
4. Additionally, HC achieved 3.8%, 7.0%, 2.1%, and 1.4% AUC performance improvements over the FasterViT [15], SHViT [16], Swin [17], and ViT [18] models, respectively. HX gained 4.2%, 7.2%, 1.6%, and 1.1% AUC performance improvements over the FasterVit, SHViT, Swin and ViT models, respectively.
5. These results demonstrate that our computational framework (1) is computationally efficient when working under the transfer learning setting; (2) leads to promoting never-ending AI model generation; (3) expands the model space from 4 to 10000; and (4) aids in the search of high-performance models with which to tackle the studied classification task.

## 2. Materials and Methods

### 2.1. Data Preprocessing

The CT dataset (named the SARS-COV-2 CT-Scan database) was downloaded from Kaggle: https://www.kaggle.com/datasets/plameneduardo/sarscov2-ctscan-dataset (accessed on 17 March 2024). We randomly selected and processed 2481 images, in which the class label distribution was 1252 images for COVID-19 and 1229 images for non-COVID-19. The X-ray dataset (named the COVID-19 Radiography Database) was downloaded from Kaggle at https://www.kaggle.com/datasets/tawsifurrahman/covid19-radiography-database (accessed on 17 March 2024). We randomly selected 2000 images, in which 1000 images were categorized as COVID-19 and the other remaining 1000 images were categorized as non-COVID-19. All images were resized to 256 × 256 when addressing the COVID-19 classification task in this study. Figure 2 illustrates sample images from the CT and X-ray datasets prior to and after processing.

**Figure 2.**
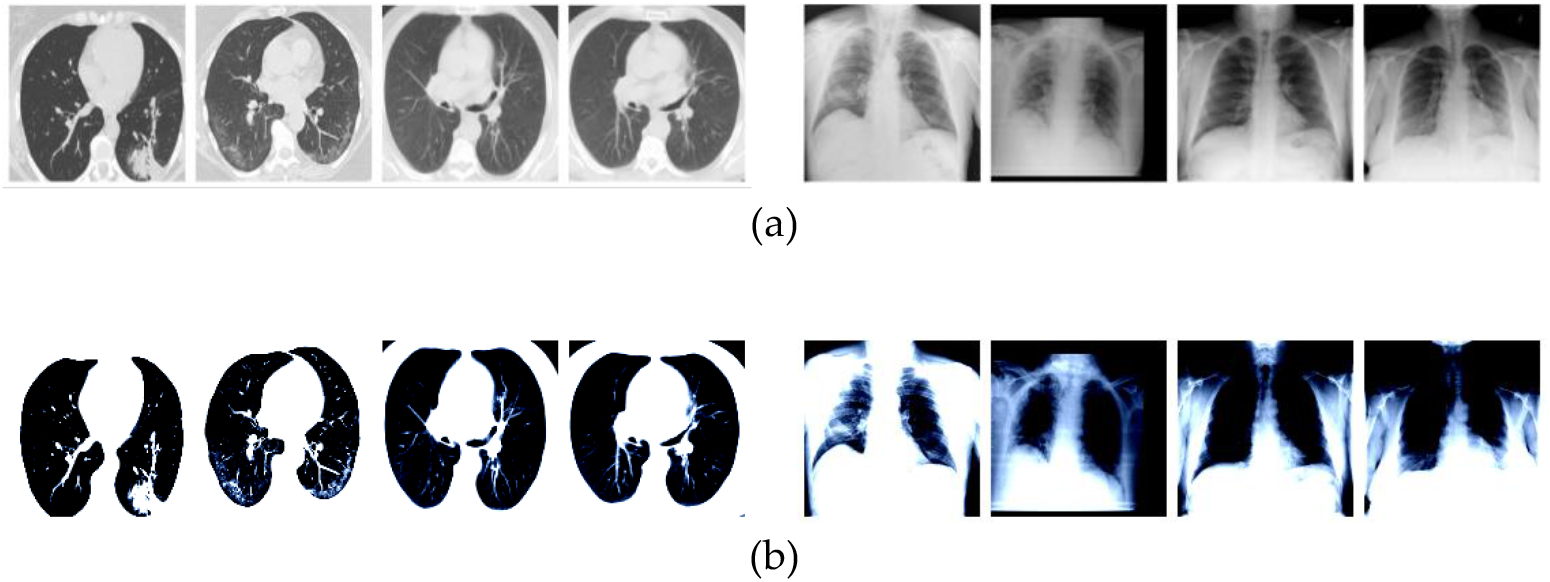
From left to right: CT and X-ray images pertaining to two COVID-19 and non-COVID-19 samples, respectively. (a) Before and (b) after processing.

### 2.2. Pre-Trained CNNs

#### 2.2.1. ConvNeXt

This pre-trained model on the ImageNet database of 1000 class labels initially starts with 1 convolutional layer, followed by 18 ConvNeXt blocks, which resemble residual blocks. However, each ConvNeXt block employs (1) layer normalization instead of batch normalization; (2) Gaussian error linear unit (GELU) activation instead of rectified linear unit (ReLU); and (3) 18 ConvNeXt blocks spanned over 4 stages as (3, 3, 9, 3) rather than 16 residual blocks spanned over 4 stages as (3, 4, 6, 3) in ResNet50. Each ConvNeXt block is composed of three convolutional layers coupled with a residual connection. Flattening is achieved before classification, which is a 1000-way fully connected dense layer coupled with a softmax activation function. This pre-trained model is referred to in [57] as ConvNeXt-T.

#### 2.2.2. EfficientNetV2

Unlike EfficientNet, which only uses MBConv blocks for feature extraction, Efficient-NetV2 [58] includes both Fused-MBConv and MBConv blocks for feature extraction. The Fused-MBConv block improves training speed via replacing the pointwise and depthwise convolutional layers in MBConv with one convolutional layer. In our study, the pretrained model initially starts with convolutional layer, followed by three Fused-MBConv blocks, three MBConv blocks, and a convolutional layer for feature extraction. A global average pooling is applied before the classification, consisting of a 1000-way fully connected dense layer coupled with a softmax activation function.

#### 2.2.3. DenseNet121

The feature extraction part in this pre-trained model [59] initially starts with one standard convolutional and pooling layer, followed by four dense blocks with three interleaved transition layers. The four dense blocks consist of 6, 12, 24, and 16 convolutional blocks. Each convolutional block is composed of two convolutional layers. The transition layers include average pooling layers for dimensionality reduction in feature maps. The classification is a 1000-way fully connected dense layer coupled with a softmax activation function.

#### 2.2.4. ResNet34

For feature extraction, the ResNet34 pre-trained model [60] on the ImageNet database of 1000 class labels initially starts with one convolutional and max pooling layer, followed by 16 residual blocks, spanned over four stages as (3, 4, 6, 3). Each residual block is composed of two convolutional layers coupled with skip connection, where batch normalization is placed after each convolution. Global average pooling is then performed before the classification, which is a 1000-way fully connected dense layer coupled with a softmax activation function.

### 2.3. DTL

In Figure 3, we illustrate the DTL part pertaining to our approach applied to CT and X-ray images. First, we utilize four pre-trained deep learning (DL) models, including ConvNeXt, EfficientNetV2, DenseNet121, and ResNet34. Each pre-trained DL model was induced after training on a million images from the ImageNet database to tackle the multiclass classification task of categorizing images into 1 of the 1000 predefined classes. We replace the 1000-way fully connected dense layer with softmax activation by training a 1-way fully connected layer with sigmoid activation. In this study, we leave the weights of the feature extraction part unchanged, and it is applied to COVID-19 images followed by a flattening step, and then training is performed to update the weights of subsequent layers in the classification part. The training phase incorporates the Adam optimizer and binary cross-entropy with a logit loss function. Each DTL model yields probabilities when performing predictions, in which, if the probability if greater than 0.5, then the image is classified as non-COVID-19. If the prediction value is less than or equal to 0.5, then the image is classified as COVID-19. In our DTL model, the fully connected dense layer is a weight layer that is not frozen and thereby has a few numbers of parameters during the training process. The only exception is ConvNeXt, in which we also train the layer normalization right before the fully connected dense layer (see Table 2).

**Table 2.** Summary for layers and parameters pertaining to the DTL models in this study.

| Model | No. of Params | Unfrozen Layers | Transferred | Trainable |
| --- | --- | --- | --- | --- |
| ConvNeXt | 27,820,897 | 2 | 27,818,592 | 2,305 |
| EfficientNetV2 | 20,178,769 | 1 | 20,177,488 | 1,281 |
| DenseNet121 | 6,954,881 | 1 | 6,953,856 | 1,025 |
| ResNet34 | 21,285,185 | 1 | 21,284,672 | 513 |

**Figure 3.**
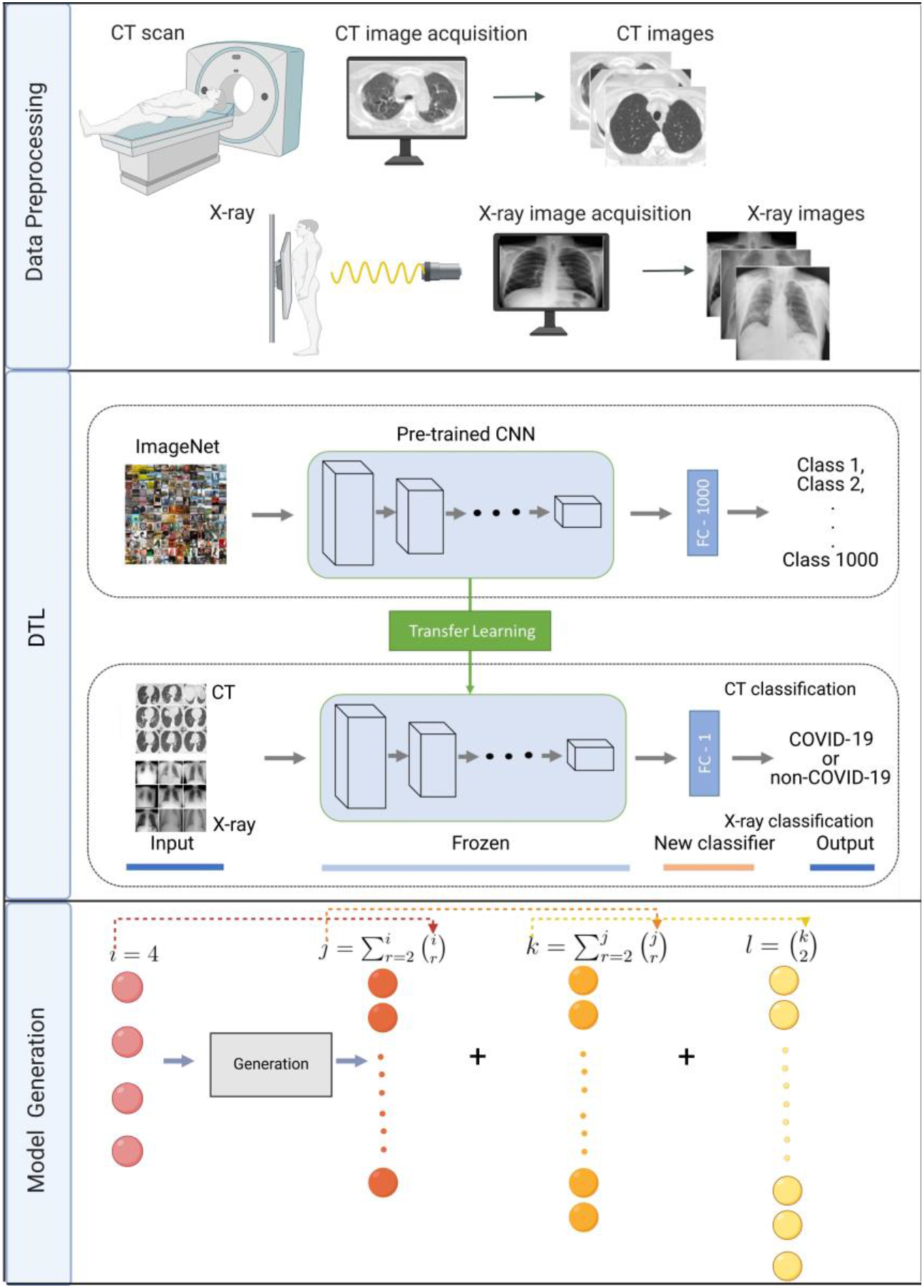
Flowchart showing our computational approach for COVID-19 classification. Data pre-processing: acquiring COVID-19 CT and X-ray images pertaining to COVID-19 and non-COVID-19 patients. DTL: CT images are provided to a pre-trained CNN for feature extraction followed by training a new classifier to discriminate between COVID-19 and non-COVID-19 images. Model generation: four models are provided as inputs to sequentially generate DTL models. Figure created with BioRender.com.

### 2.4. Model Generation

To generate a large number of DTL models performing prediction for a test example, *z*, in our study, we start with 4 DTL models, namely *h*_1_(*z*), *h*_2_(*z*), *h*_3_(*z*), and *h*_4_(*z*), which correspond to models obtained from training ConvNeXt, EfficientNetV2, DenseNet121, and ResNet34, respectively. Let D1={*h*_1_(*z*), *h*_2_(*z*), *h*_3_(*z*), *h*_4_(*z*)}, where *hi*(*z*): *z*→{COVID-19, non-COVID-19}, and we generate all 2, 3 and 4 combinaions from D1, resulting in 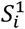 for *i=*1…11 attributed to 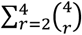. It can be seen that the size of a given 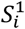 is upper bounded by 4 and lower bounded by 2 (i.e., 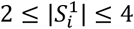). Assuming that 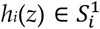, we then perform majority prediction as shown in Equation 1, in which *I*() is an indicator function returning 1 if true and 0 otherwise. Let D2={*h*1_1_(*z*), *h*1_2_(*z*),…, *h*1_11_(*z*)}. Now, we generate all 2, 3, 4, 5, 6, 7, 8, 9, 10, and 11 combinations from D2, resulting in 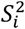 for *i* = 1…2036 ascribed to 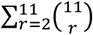. It can be seen that the size of a given 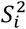 is lower bounded by 2 and upper bounded by 11 (i.e.,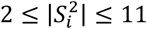). Let 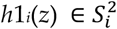 prediction is performed using Equation 2. We set D3 = {*h*2_1_(*z*), *h*2_2_(*z*),…, *h*2_2036_(*z*)}. We only select 7953 from the generated 2 combinations of D3. Therefore, we have 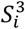 for *i* = 1…7953 from 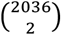 models. For 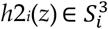, prediction is performed using Equation 3. It can be shown that we had 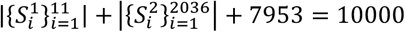 models.

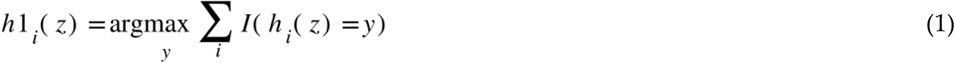

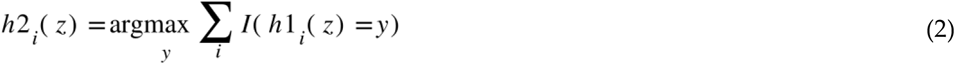

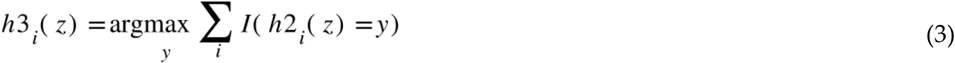

### 2.5. Classification Methodology

In our study, we randomly adapted four pre-trained DL models, including ConvNeXt, EfficientNetV2, DenseNet121, and ResNet34. All of these four pre-trained DL models were trained on more than a million images from the ImageNet database, in which the number of different class labels is equal to 1000. We froze all layers in feature extraction related to convolutional and pooling layers to extract features from the COVID-19 images generated via imaging techniques while updating weights in the subsequent layers pertaining to the classification part during the training phase. For all of the DTL models, we utilized the Adam optimizer with binary cross-entropy with a logit loss function. In terms of the optimization parameters when building models during the training phase, we used 32 for the batch size, 0.001 for the learning rate, and 10 for the number of epochs. When performing prediction for an unseen image, if the predicted value is greater than 0.5, then we designate the image as non-COVID-19. Otherwise, we designate the image as COVID-19. For all of the DTL models including the 10000 constructed models in each imaging technique, we employed the following five performance measures to evaluate the studied models [61, 62]:

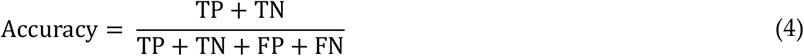

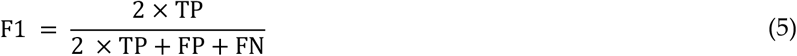

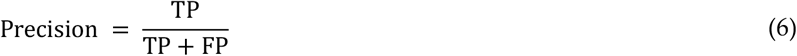

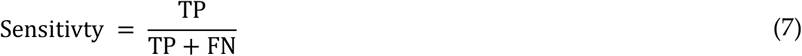

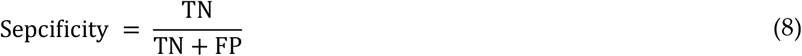

where TP stands for true positive, referring to the number of non-COVID-19 images that were correctly classified as non-COVID-19. FN stands for false negative, referring to the number of non-COVID-19 images that were incorrectly classified as COVID-19. TN stands for true negative, referring to the number of COVID-19 images that were correctly classified as COVID-19. FP stands for false positive, referring to the number of COVID-19 images that were incorrectly classified as non-COVID-19. To assess the performance on whole datasets, we employed five-fold cross-validation.

### 2.6. Implementation Details

When running the experiments in this study using an NVIDIA Tesla P100 GPU, the deep learning models are implemented in Python (version 3.10.13) [63] based on the PyTorch library (version 2.1.2) [64, 65] and the torchvision library (version 0.16.2), in which the installed CUDA version is 12.1 [66]. Data processing is achieved through various libraries, including NumPy (version 1.26.4) and pandas (version 2.2.2) [67, 68]. The performance evaluation is performed through the sklearn library (version 1.2.2) [69].

## 3. Results

### 3.1. Classification Results

#### 3.1.1. Training Results

Figure 4a and b display the average training accuracy and loss over 10 epochs for the eight DTL models during the running of five-fold cross-validation, obtained when applied to COVID-19 CT and X-ray datasets, respectively. The accuracy and loss moving in the upward and downward directions, respectively, show the capability of models to learn and thereby will be able to classify unseen COVID-19 images during the testing phase. When considering the CT dataset (Figure 4a), ViT achieved the highest and lowest average accuracy and loss of 0.871 and 0.257, respectively, followed by ConvNeXt, achieving an average accuracy (and loss) of 0.860 (and 0.260). DenseNet121, yielding an average accuracy (and loss) of 0.846 (and 0.303). SHViT had the lowest average accuracy (and a loss) of 0.806 (and 0.356). For the X-ray dataset as shown in Figure 4b, ConvNeXt achieved the highest average accuracy of 0.899 while attaining the lowest average loss of 0.204. ViT had an average accuracy and loss of 0.893 and 0.215, respectively. Swin obtained an average accuracy (and loss) of 0.877 (and 0.230). DenseNet121 achieved an average accuracy and loss of 0.870 and 0.260, respectively. SHViT yielded an average accuracy and loss of 0.831 and 0.318, respectively.

**Figure 4.**
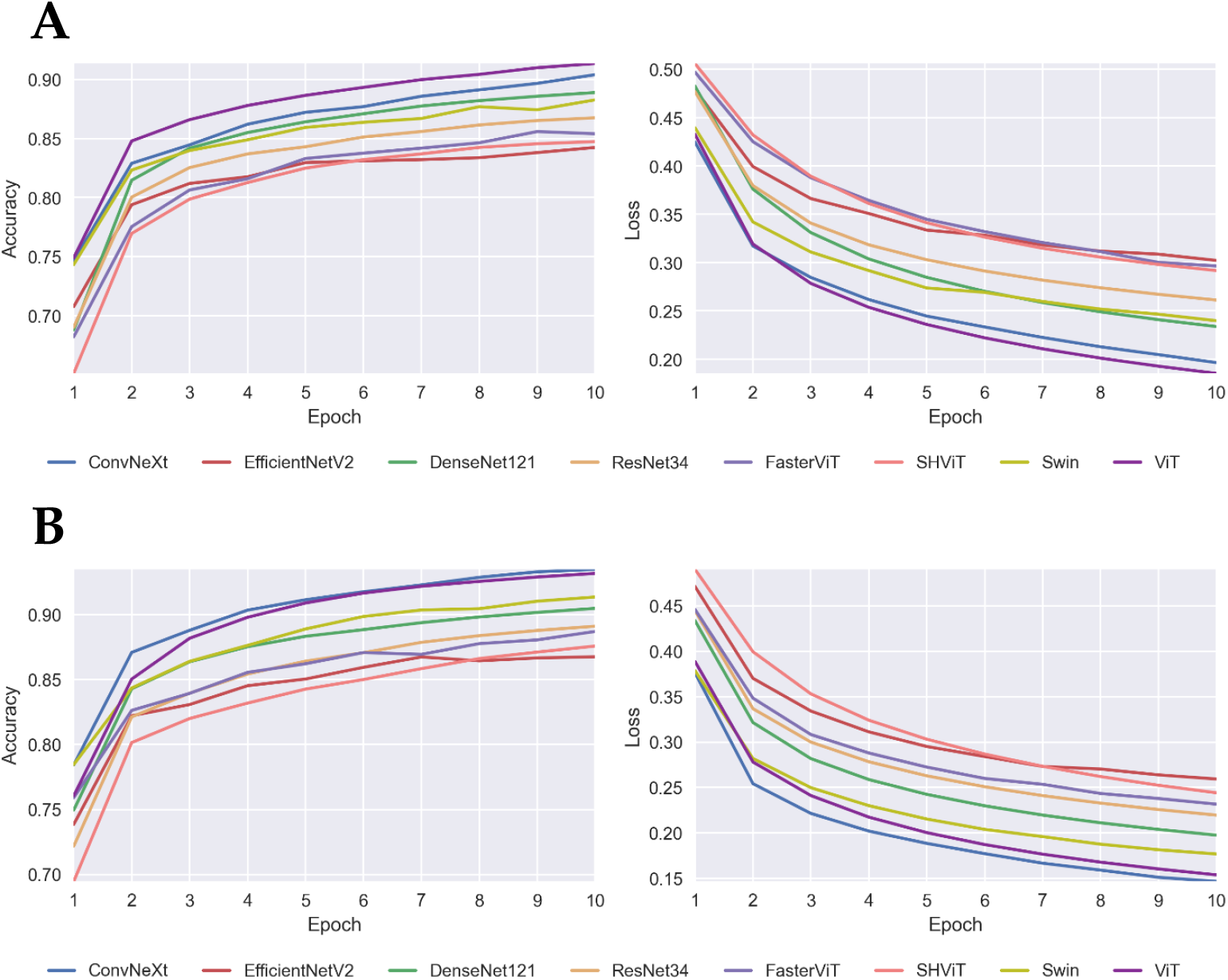
Average accuracy and loss for ten epochs during the running of five-fold cross-validation pertaining to (a) CT and (b) X-ray datasets.

In Figure 5a and b pertaining to CT and X-ray datasets, respectively, we illustrate the total running time for the model induction process of four DTL models during the running of five-fold cross-validation. In terms of the CT dataset as shown in Figure 5a, ResNet34 is the fastest. Specifically, ResNet34 is 1.14 × faster than ConvNeXt. Moreover, the running time difference between DenseNet121 and EfficientNetV2 is marginal, in which the former and latter are 1.067 and 1.062 × faster than ConvNeXt. For the X-ray dataset as illustrated in Figure 5b, ResNet34 is still the fastest method, which is 1.37 × faster than ConvNeXt. The second and third fastest methods are DenseNet121 and EfficientNetV2, respectively. Both DenseNet121 and EfficientNetV2 resulted in marginal running time differences in which the former and latter are 1.242 and 1.210 × faster than ConvNeXt. These results demonstrate that, although ConvNeXt yielded the highest performance results, it was the slowest among all of the models. Because we did not incorporate transformer-based models in the model generation process, we moved the total running time of each transformer-based model in Figure S3 within the Supplementary Additional File.

**Figure 5.**
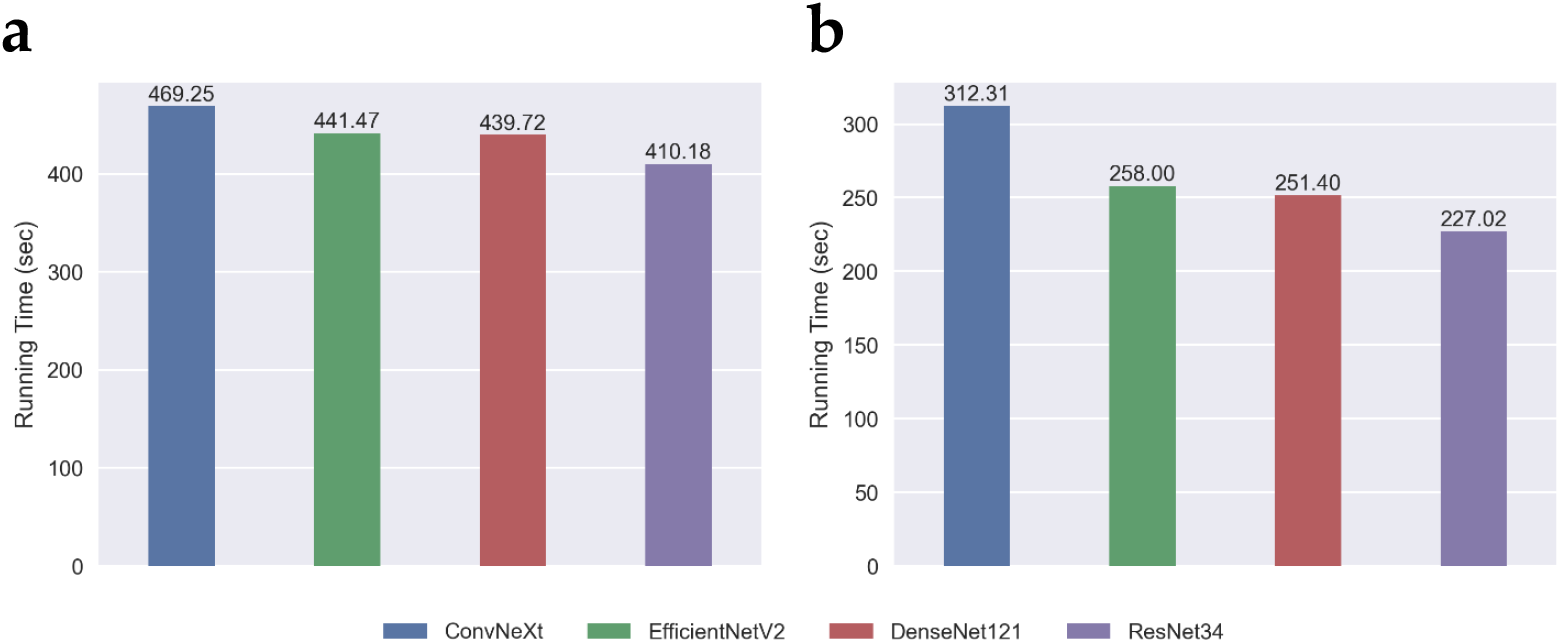
Total running time for the four studied DTL models during the training phase in five-fold cross-validation pertaining to (a) CT and (b) X-ray datasets.

#### 3.1.2. Testing Results

The boxplots in Figure 6a and b illustrate the generalization (testing) results of the four DTL models utilizing five-fold cross-validation pertaining to CT and X-ray datasets, respectively. Regarding Figure 6a and b, the median results are indicated via the horizontal lines that cross each box. In terms of the CT dataset in Figure 6a, ConvNeXt yielded the best results, achieving the highest medians of 0.901, 0.903, 0.874, 0.935, and 0.872 in terms of accuracy, F1, precision, sensitivity, and specificity, respectively. The second best performing model was DenseNet121, achieving a median accuracy of 0.885, a median F1 of 0.889, a median precision of 0.857, a median sensitivity of 0.930, and a median specificity of 0.844. ResNet34 is the third best performing model, generating a median accuracy of 0.853, a median F1 of 0.863, a median precision of 0.832, a median sensitivity of 0.894, and a median specificity of 0.828. EfficientNetV2 is the worst performing method, yielding the lowest median accuracy of 0.842, the lowest median F1 of 0.851, the lowest median precision of 0.802, a median sensitivity of 0.910, and the lowest median specificity of 0.780. Regarding the X-ray dataset in Figure 6b, ConvNeXt obtained the highest median accuracy of 0.930, the highest median F1 of 0.928, the highest median precision of 0.931, the highest median sensitivity of 0.930, and the highest median specificity of 0.930. The second best performing method is DenseNet121, which had a median accuracy of 0.917, a median F1 of 0.912, a median precision of 0.910, a median sensitivity of 0.925, and a median specificity of 0.915. The third best method is ResNet34, yielding a median accuracy, F1, precision, sensitivity, and specificity of 0.885, 0.887, 0.896, 0.900, and 0.895, respectively. EfficientNetV2 was the worst performing model, obtained a median accuracy, F1, precision, sensitivity, and specificity of 0.865, 0.862, 0.848, 0.915, and 0.835, respectively.

**Figure 6.**
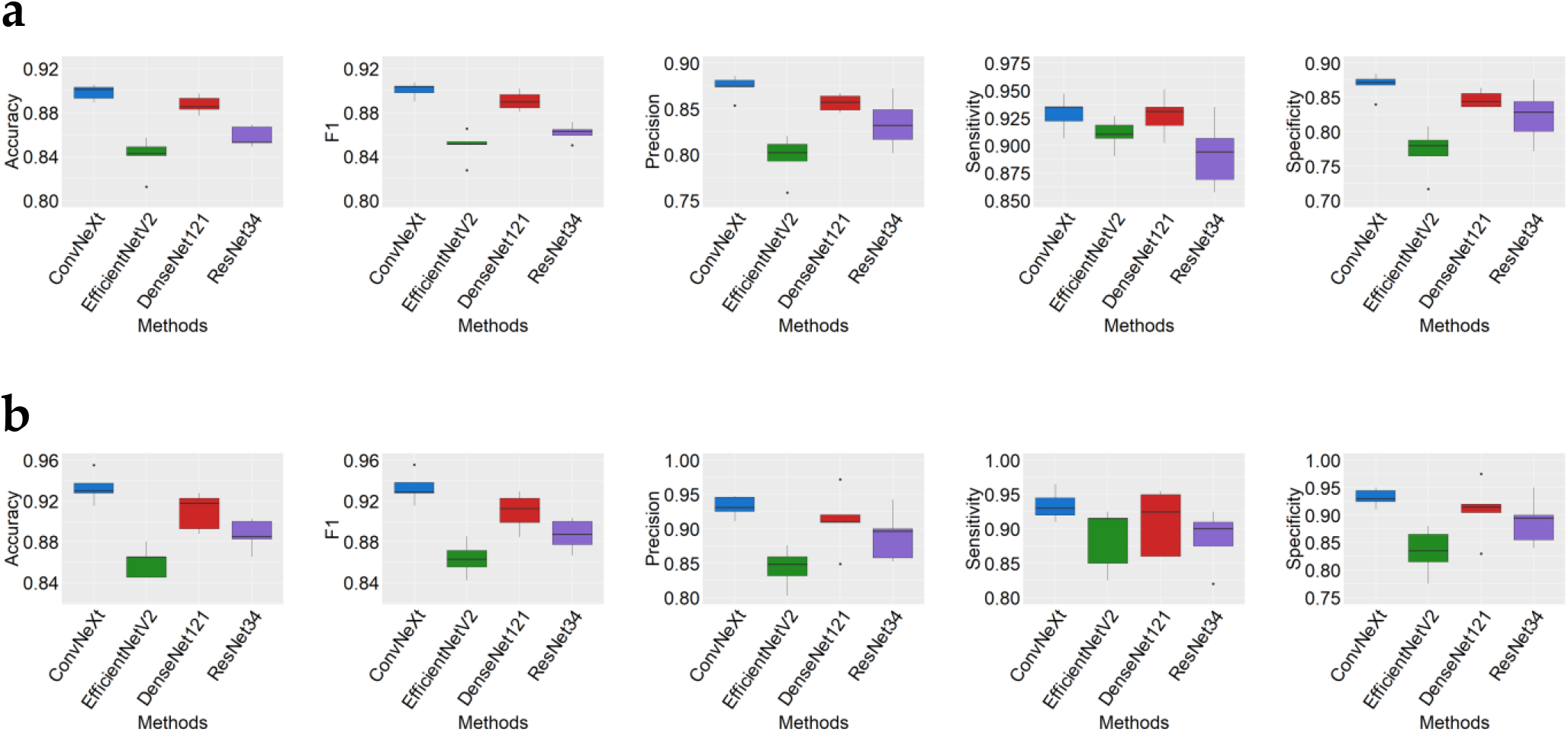
Boxplots illustrate several generalization (testing) performance results (based on accuracy, F1, precision, sensitivity, and specificity) of the four DTL models when utilizing five-fold cross-validation pertaining to (a) CT and (b) X-ray datasets.

Table 3a and b report the testing performance results on the whole datasets pertaining to (a) CT and (b) X-ray datasets, including four transformer-based models. In terms of the CT dataset (see Table 3a), ConvNeXt is the best performing model, yielding the highest ACC of 0.894 (tie with ViT) and the highest F1 of 0.900. The second best model is ViT, which yielded an ACC of 0.894, an F1 of 0.896, the highest PRE of 0.877, an SEN of 0.915, and the highest SPE of 0.874. SHViT is the worst performing model, yielding the lowest ACC of 0.839, the lowest F1 of 0.843, a PRE of 0.813, the lowest SEN of 0.876, and an SPE of 0.802. When considering the X-ray dataset as shown in Table 3b, ConvNeXt is still the best performing model, achieving the highest ACC, F1, PRE, SEN, and SPE of 0.933, 0.933, 0.933, 0.934, and 0.932, respectively. ViT, the second best performing model, obtained an ACC, F1, PRE, SEN, and SPE of 0.919, 0.918, 0.929, 0.907, and 0.931, respectively. The worst performing model, SHViT, obtained the lowest ACC, F1, and SEN of 0.858, 0.855, and 0.839, respectively. These reported performance results illustrate the high-performance results of ConvNeXt.

**Table 3.**
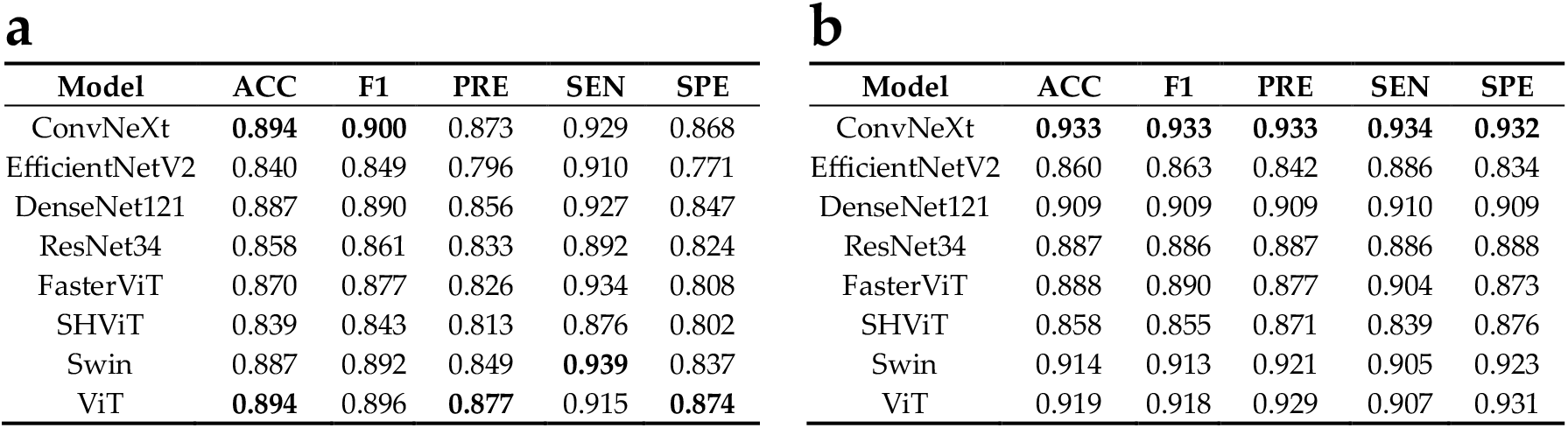
Generalization (testing) results of the eight DTL models pertaining to (a) CT and (b) X-ray datasets. ACC is accuracy. PRE is precision. SEN is sensitivity. SPE is specificity. Bold indicates a model obtaining the highest performance results.

| **a** |  |  |  |  |  | **b** |  |  |  |  |  |
| --- | --- | --- | --- | --- | --- | --- | --- | --- | --- | --- | --- |
| Model | ACC | F1 | PRE | SEN | SPE | Model | ACC | F1 | PRE | SEN | SPE |
| ConvNeXt | **0.894** | **0.900** | 0.873 | 0.929 | 0.868 | ConvNeXt | **0.933** | **0.933** | **0.933** | **0.934** | **0.932** |
| EfficientNetV2 | 0.840 | 0.849 | 0.796 | 0.910 | 0.771 | EfficientNetV2 | 0.860 | 0.863 | 0.842 | 0.886 | 0.834 |
| DenseNet121 | 0.887 | 0.890 | 0.856 | 0.927 | 0.847 | DenseNet121 | 0.909 | 0.909 | 0.909 | 0.910 | 0.909 |
| ResNet34 | 0.858 | 0.861 | 0.833 | 0.892 | 0.824 | ResNet34 | 0.887 | 0.886 | 0.887 | 0.886 | 0.888 |
| FasterViT | 0.870 | 0.877 | 0.826 | 0.934 | 0.808 | FasterViT | 0.888 | 0.890 | 0.877 | 0.904 | 0.873 |
| SHViT | 0.839 | 0.843 | 0.813 | 0.876 | 0.802 | SHViT | 0.858 | 0.855 | 0.871 | 0.839 | 0.876 |
| Swin | 0.887 | 0.892 | 0.849 | **0.939** | 0.837 | Swin | 0.914 | 0.913 | 0.921 | 0.905 | 0.923 |
| ViT | **0.894** | 0.896 | **0.877** | 0.915 | **0.874** | ViT | 0.919 | 0.918 | 0.929 | 0.907 | 0.931 |

Figure 7a and b unveil computational insights for the generalization capability of the four studied DTL models, applied to testing images pertaining to CT and X-ray datasets, respectively. Prediction differences for ConvNeXt (and all other models) between COVID-19 and non-COVID-19 images were statistically significant (*P* < 2.2 × 10^−16^ from a *t*-test), indicating that all the models are general predictors for COVID-19 or non-COVID-19 image prediction. Similarly, for the X-ray dataset in Figure 7b, prediction differences in all models were statistically significant (*P* < 2.2 × 10^−16^ from a *t*-test), demonstrating that these models are general, rather than specific predictors for COVID-19 prediction.

**Figure 7.**
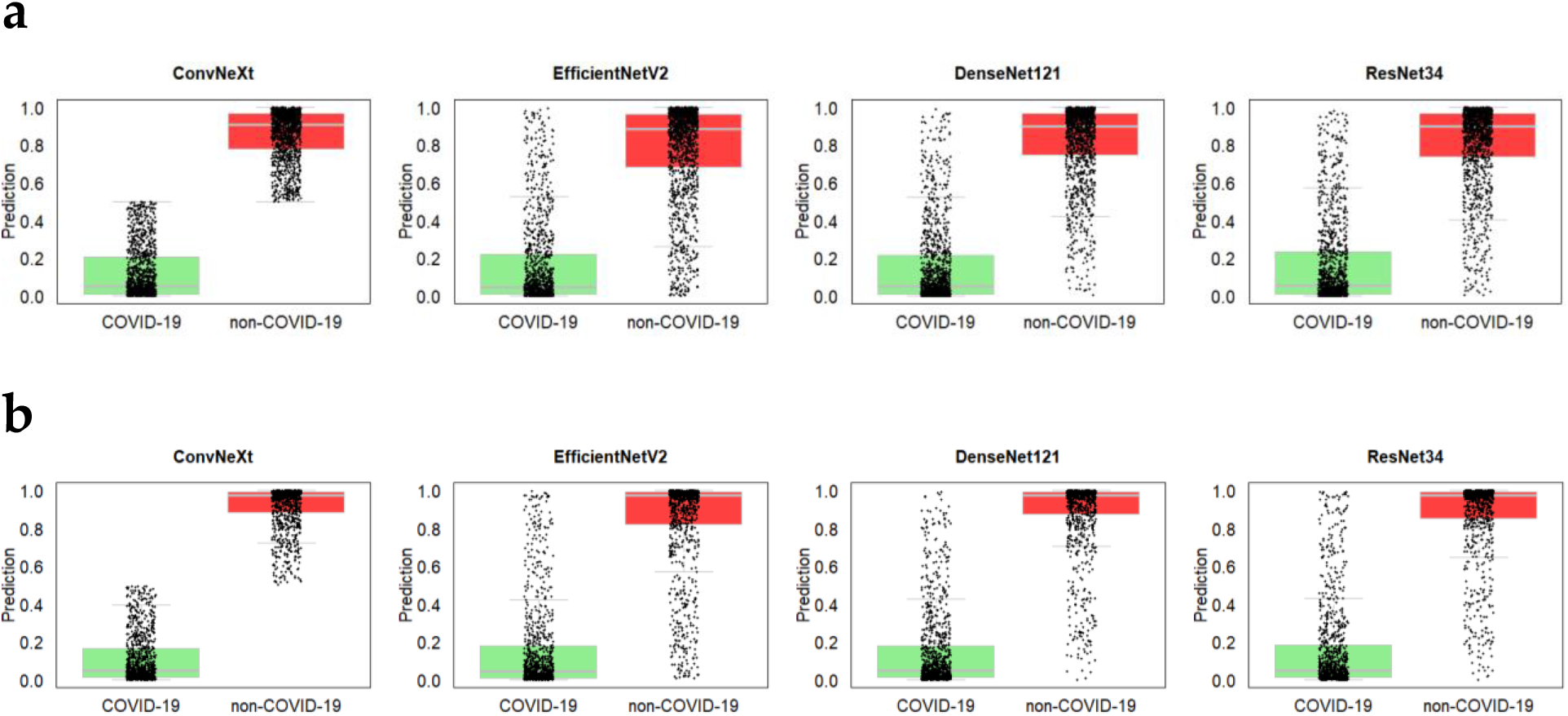
Boxplots and strip charts of predicted COVID-19 and non-COVID-19 images on all testing folds of the four DTL models pertaining to (a) CT and (b) X-ray datasets.

After carrying out the model generation process explained in Section 2.4., we obtained 10000 models and plotted their AUC results when applied to CT and X-ray datasets, as displayed in Figure 8a and b, respectively. For the CT dataset in Figure 8a, we selected the best model, which we named HC, constructed as follows: Let *h*_1_(z), *h*_2_(z), *h*_3_(z), and *h*_4_(z) correspond to the DTL models obtained from training ConvNeXt, EfficientNetV2, DenseNet121, and ResNet34, respectively, and applied to an unseen image, *z*. In Equation 9, *h*1_1_(z) corresponds to a majority vote pertaining to two classifiers, namely ConvNeXt and EfficientNetV2, applied to a testing example. *I* is an indicator function resulting in 1 if the arguments are true and 0 otherwise. *h*1_2_(z) in Equation 10 corresponds to the majority vote of two classifiers (i.e., ConvNeXt and DenseNet121). In Equation 11, *h*1_9_(z) corresponds to a majority vote of three classifiers (i.e., ConvNeXt, DenseNet121, and ResNet34). HC in Equation 12 is constructed as a majority vote mechanism of *h*1_1_(z), *h*1_2_(z), and *h*1_9_(z). Figure S1 in the Supplementary Additional File provides an additional illustration for HC.

**Figure 8.**
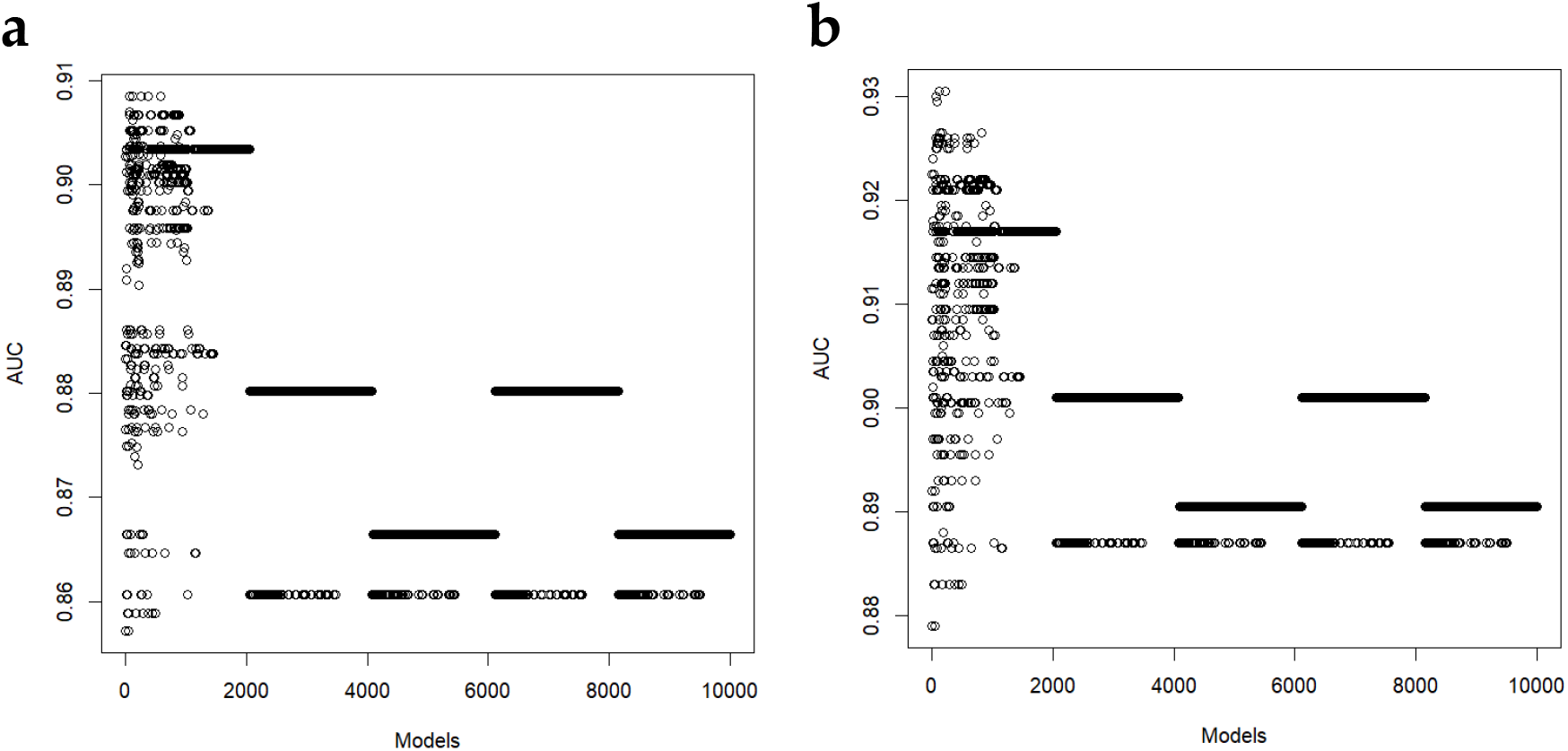
AUC performance results of 10000 generated models pertaining to (a) CT and (b) X-ray datasets.

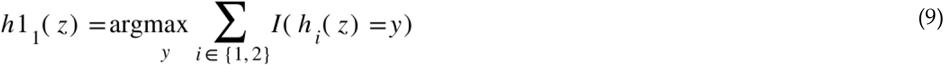

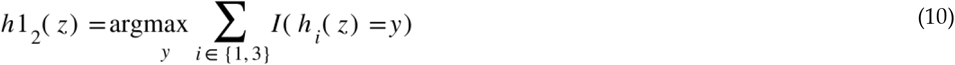

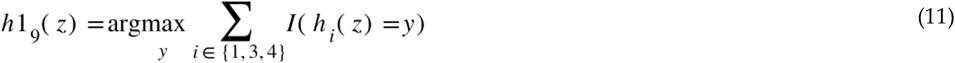

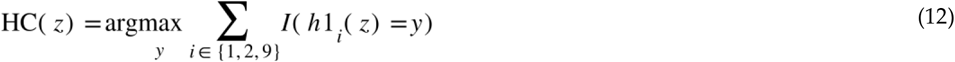

The AUC for HC is 0.909 as illustrated in Figure 9a. For the X-ray dataset as illustrated in Figure 8b, we selected the best model attaining the highest AUC performance and designated it HX, working as shown in Equations 13–16 as follows: *h*1_2_(z) in Equation 13 corresponds to a majority vote pertaining to two classifiers (i.e., ConvNeXt and DenseNet121). *h*1_3_(z) in Equation 14 is the majority vote of ConvNeXt and ResNet34. *h*1_9_(z) in Equation 15 is just the majority vote of ConvNeXt, Dense-Net121, and ResNet34. In Equation 16, HX is the majority vote of *h*1_2_(z), *h*1_3_(z), and *h*1_9_(z). We report the obtained AUC results via HX in Figure 9b. Figure S2 in the Supplementary Additional File provides an additional illustration for HX.

**Figure 9.**
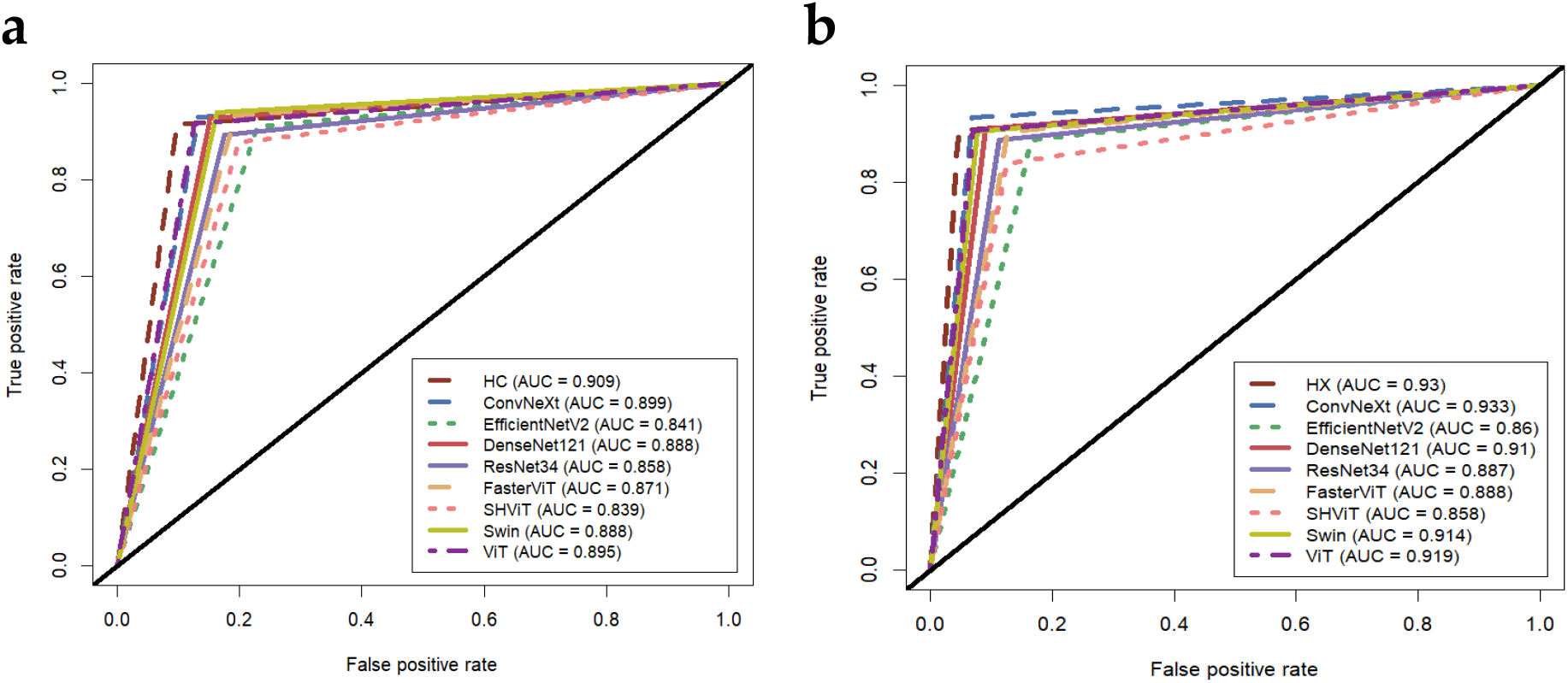
ROC curves and recorded AUC performance results of eight DTL models against the generated models HC and HX to (a) CT and (b) X-ray datasets.

Compared to transformer-based models including FasterViT, SHViT, Swin, and ViT using the CT dataset (see Figure 9a), HC achieves 3.8%, 7.0%, 2.1%, and 1.4% AUC performance improvements over the FasterViT, SHViT, Swin and ViT models, respectively. ConvNeXt yields 2.8%, 6.0%, 1.1%, 0.4% AUC performance improvements over FasterViT, SHViT, Swin, and ViT, respectively. When considering the X-ray dataset as shown in Figure 9b, HX yields 4.2%, 7.2%, 1.6%, and 1.1% AUC performance improvements over the FasterVit, SHViT, Swin and ViT models, repressively. ConvNeXt generates 4.5%, 7.5%, 1.9%, and 1.4% AUC performance improvements over FasterViT, SHViT, Swin, and ViT, respectively.

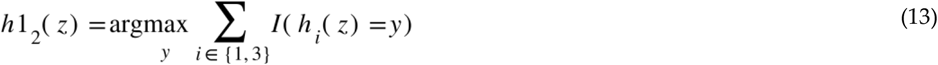

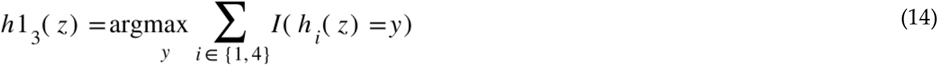

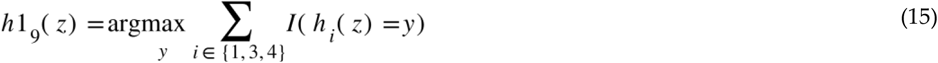

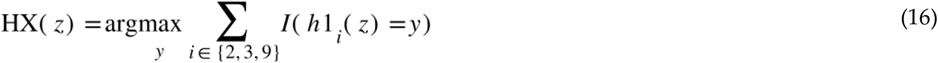

Figure 10a and b report the confusion matrices calculated based on testing for CT and X-ray datasets, respectively. When considering the CT dataset, it can be shown that HC is the best model, accurately predicting 2254 out of 2481 CT images. One hundred and twenty-one images were predicted as non-COVID-19 while their actual label was COVID-19, and thereby counted as one hundred and twenty-one FP. One hundred and six images were predicted as COVID-19 while their actual label was non-COVID-19, therefore counted as one hundred and six FN.

**Figure 10.**
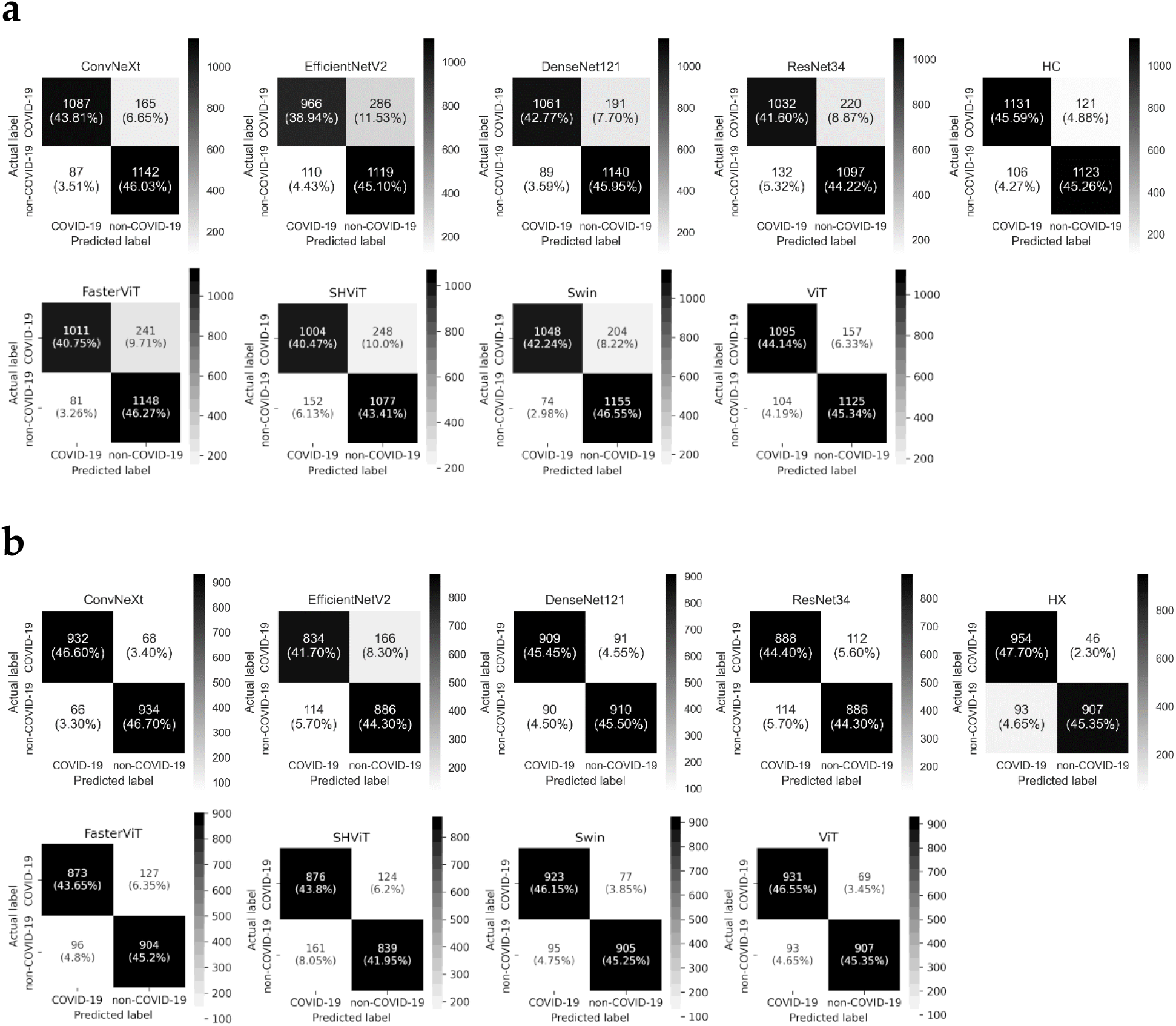
The combined confusion matrices of five testing folds during the running of five-fold cross-validation for ten models pertaining to (a) CT and (b) X-ray datasets.

The second best model is ConvNeXt, accurately predicting 2229 out of 2481 CT images. One hundred and sixty-five images were predicted by ConvNeXt as non-COVID-19 while their actual label was COVID-19, and thereby counted as one hundred and sixty-five FP. Eighty-seven images were incorrectly predicted as COVID-19 while their actual label was non-COVID-19, therefore counted as eighty-seven FN. For the X-ray images, ConvNeXt accurately predicts 1866 (934 TP + 932 TN) images, which is marginally higher than HX (1861 calculated from 907 TP + 954 TN). Both ConvNeXt and HX inaccurately predict 134 (68 FP + 66 FN) and 139 (46 FP + 93 FN) images, respectively.

In terms of transformer-based models (i.e., FasterViT, SHViT, Swin, and ViT) when using the CT dataset, the combined confusion matrices of Figure 10a demonstrate that FasterViT, SHViT, Swin, and ViT accurately predict 2159, 2081, 2203, and 2220 images, respectively, out of 2481 CT images. HC, which accurately predicted 2254 CT images (see Figure 10a), had 95, 173, 51, and 34 more accurately predicted images than FasterViT, SHViT, Swin, and ViT, respectively. FasterViT, SHViT, Swin, and ViT had 241, 248, 204, and 157 images, respectively, predicted as non-COVID-19 while their actual label was COVID-19, and thereby counted as 241 FP, 248 FP, 204 FP, and 157 FP (compared to 121 FP for HC), respectively. Moreover, FasterViT, SHViT, Swin, and ViT had 81, 152, 74, and 104 images, respectively, predicted as COVID-19 while their actual label was non-COVID-19, and thereby counted as 81 FN, 152 FN, 74 FN and 104 FN (compared to 106 FN for HC), respectively. For the X-ray dataset as shown in Figure 10b, FasterViT, SHViT, Swin, and ViT had a total of 1777, 1715, 1828 and 1838 accurately predicted images, respectively, out of 2000 (compared to 1861 for HX as shown in Figure 10b). Hence, HX had 84, 146, 33, and 23 more accurately predicted images than FasterViT, SHViT, Swin, and ViT, respectively. FasterViT, SHViT, Swin, and ViT had 127, 124, 77, and 69 images, respectively, that were predicted as non-COVID-19 while their actual label was COVID-19; therefore, they had 127 FP, 124 FP, 77 FP, and 69 FP (compared to 46 FP for HX), respectively. FasterViT, SHViT, Swin, and ViT had 96 FN, 161 FN, 95 FN, and 93 FN, respectively, compared to 93 FN for HX.

Figure 11 provides a summary for the number of updated parameters during the training process. FasterViT (tie with ResNet34) has the least number of parameters when compared to all other models including transformer-based models (SHViT, Swin, and ViT). Additional details about layers and number of parameters pertaining to transformer-based models are provided in Supplementary Additional File (see Table S1).

**Figure 11.**
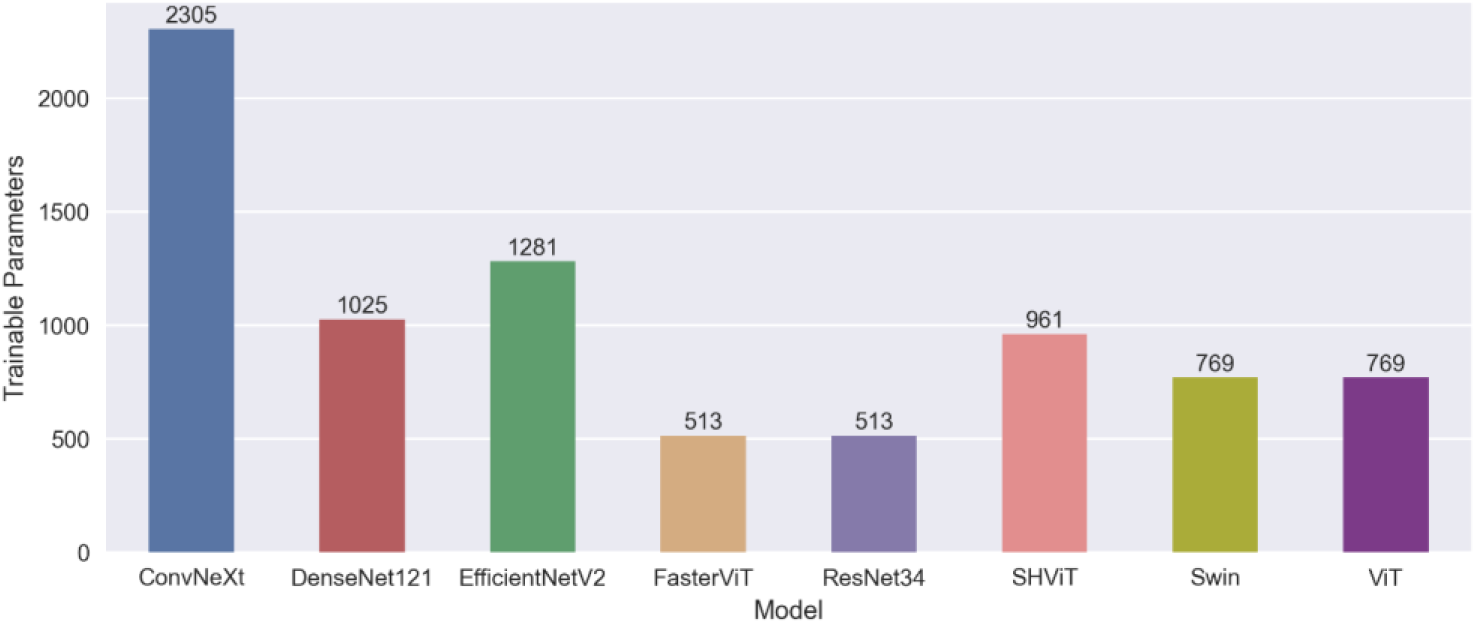
An overview for the number of updated parameters pertaining to each studied model.

## 4. Discussion

To conduct a large empirical study on COVID-19 classification using CT and X-ray images, our computational approach comprises three parts: data preprocessing and DTL followed by model generation. In terms of the data preprocessing part, CT images composed of 2481 images pertaining to COVID-19 and non-COVID-19 patients were down-loaded from Kaggle at https://www.kaggle.com/datasets/plameneduardo/sarscov2-ctscan-dataset. The 2000 X-ray images regarding COVID-19 and non-COVID-19 were downloaded from Kaggle at https://www.kaggle.com/tawsifurrahman/covid19-radiography-database. We prepared all PNG images to be of size 256 × 256. For the DTL computation, we employed four pre-trained DL models, including ConvNeXt, Efficient-NetV2, DenseNet121, and ResNet34. Each of these DTL models was already trained on more than a million images from the ImageNet database. We extracted features from

COVID-19 images using weights obtained from pre-trained models on the ImageNet database, followed by a flattening step before training from scratch the densely connected classifier with the Adam optimizer and binary cross-entropy with a logit loss function. The model generation part receives four DTL models. Then, taking the majority of two, three, and four combinations from the four DTL models resulted in 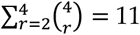 constructed models. After that, taking the majority of 2, 3, …, 11-combinations from the 11 models resulted in 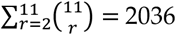 constructed models. Finally, we selected 7953 models from 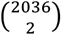.

In our computational approach, it can be seen from Equations 1–3 that *h*3_i_ depends on *h*2_i_, which also depends on *h*1_i_. Therefore, it can be viewed as *h*3_i_(*h*2_i_(*h*1_i_(z))), and this demonstrates the sequential nature for constructing these generated models. For the HC model as shown in Equations 9–12, Equations 9, 10, and 11 are exactly like calculations in Equation 1, while HC in Equation 12 is like *h*2_i_ in Equation 2. Hence, it can be viewed as HC(*h*1_1,2,9_(z)). The same computational nature holds true for HX corresponding to HX(*h*1_2,3,9_(z)). Figure 8 reports the AUC for 10000 models applied to CT and X-ray datasets, respectively. In Supplementary ctLGM and xrayLGM, we report the 10000 constructed models obtained from the CT and X-ray datasets, respectively.

To ensure the fairness of the experimental comparisons, we utilized five-fold cross-validation in which in all of the five runs the training is used to induce models while testing is used to perform prediction and evaluation. The sum of entries of each confusion matrix in Figure 10 is equal to the number of images in the studied datasets. The sum of each confusion matrix in Figure 10a is equal to 2481. Similarly, the sum of entries in each confusion matrix in Figure 10b is equal to 2000. This demonstrates that we evaluated the performance on the studied datasets. The generalization performance is based on evaluating the prediction performance of unseen images against their ground truth, and thereby, it is achieved in a try-and-see manner. Additionally, DTL models in our study are computationally efficient, attributed to few updated weight layers in the classification layer during the training process.

In terms of reducing the overfitting effect for induced DTL models, we included a dropout layer in the classification part pertaining to the densely connected classifier, setting the ratio to 0.2. Furthermore, we transferred weights in the feature extraction part of pre-trained models already trained on more than one million images while training from scratch the classification part in which weights in those layers are changed. In other words, incorporating a dropout layer as well as transfer learning via freezing all layers in the feature extraction part contributed to the mitigation of overfitting in this study [70].

Table 1 provides an overview of 20 existing studies, showing that our proposed study is unique when considering the number of models (see the no. of models column). There is no large empirical DL study to date, to the best of our knowledge, reporting and investigating the feasibility of DL when tackling the COVID-19 classification task using CT and X-ray images like ours. Specifically, for each imaging technique (i.e., CT or X-ray), we reported the results of eight DTL models plus 10000 generated DTL models (see Figure 8). These results show the power of our computational approach, and we have 10008 reported results for CT images + 10008 reported results for X-ray images, resulting in a total of 20016 reported results. For the conducted experiments, we did not perform data augmentation while we normalized the images with a specific mean and standard deviation using the normalize function in the torchvision library. Moreover, we found that a batch size of 32 with 10 epochs works better in our experiments with the small datasets at hand than a larger number of epochs. Hence, we halted the training after 10 epochs.

Our computational framework has both advantages and disadvantages. Some advantages include (1) the ability to expand the model space in our study to thousands of models in less than two hours, attributed to fast training time under the transfer learning setting, and (2) the availability of a large number of models that can be exploited via clustering models of similar performance differences for use in learning scenarios such as active learning. However, our framework requires additional training time in cases demanding large DL models obtained via training from scratch in which scaling to thousands of models becomes more expensive.

We employed the majority vote mechanism within the performed combinations to create new models and thereby expanding the model space, potentially identifying a better performing model. These constructed models can aid in the search for better performance models in which more automatically constructed models increase the odds for identifying a better performing model, as reported in our experimental results. Moreover, constructed models can be useful in active learning strategies such as the committee construction in the query by committee [71, 72].

## 5. Conclusions and Future Work

In this work, we presented a novel computational approach with which to augment and expand empirical studies for the task of COVID-19 classification using CT and X-ray images. First, we downloaded and processed 2481 CT and 2000 X-ray images from Kaggle for the COVID-19 binary classification task. Then, we provided processed images of each imaging technique to four pre-trained DL models, including ConvNeXt, EfficientNetV2, DenseNet121, and ResNet34. To employ transfer learning, we froze all layers, including convolutional and pooling layers, in the feature extraction part applied to COVID-19 images while unfreezing and updating weights during the training phase in the classification part of the densely connected classifier using the Adam optimizer and binary cross-entropy with a logit loss function. The generation of models was achieved in a sequential manner, generating 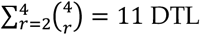 models through taking the majority vote of two, three, and four combinations from the four DTL models, and then generated 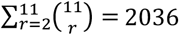 DTL models through taking majority vote of 2, 3, …, 11 combinations from 11 DTL models. Finally, 7953 DTL models were generated from 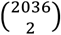. For each imaging technique, we report results for 10004 (4 + 11 + 2036 + 7953) DTL models plus 4 transformer-based models. Generalization (testing) results demonstrate that a generated DTL model, named HC, generated the highest AUC of 0.909 when applied to COVID-19 CT images. For the COVID-19 X-ray images, ConvNeXt generated the highest AUC of 0.933, which is marginally higher than the generated DTL, named HX, yielding an AUC of 0.930. The results attributed to the computational approach successfully demonstrate how to scale empirical studies as well as set a unique record for COVID-19 classification.

Future work can include the following: (1) adopting the presented computational approach to generate and establish a committee of models to be utilized in learning algorithms under an active learning scenario; (2) adjusting the computational approach to construct models via interleaved weighted majority vote; (3) expanding the model generation to reach one million constructed models; and (4) employing the presented approach to augment and enlarge empirical studies for other problems in biology and medicine.

## Data Availability

The CT dataset in this study is available at https://www.kaggle.com/datasets/plamenedu-ardo/sarscov2-ctscan-dataset, accessed on 17 March 2024. The X-ray dataset is available at https://www.kaggle.com/datasets/tawsifurrahman/covid19-radiography-database, accessed on 17 March 2024.

## Author Contributions

T.T. conceived and designed the study. M.A. performed deep learning experiments. T.T. and M.A. contributed to the visualization of results. T.T., Y-h.T., and M.A. evaluated the results, discussions, and outcomes, and wrote and reviewed the manuscript. All authors have read and agreed to the submitted version of the manuscript.

## Abbreviations

The following abbreviations are used in this manuscript:

DL: Deep learning
DTL: Deep transfer learning
LGM: Large generation of models
DenseNet: Dense convolutional network
ResNet: Residual neural network
Swin: Shifted windows
ViT: Vision transformer
SHViT: Single-head vision transformer
CNN: Convolutional neural network
SARS-CoV-2: Severe acute respiratory syndrome coronavirus 2
COVID-19: Coronavirus disease 19
Adam: Adaptive moment estimation
CT: Computed tomography
ACC: Accuracy
AUC: Area under the ROC curve
PRE: Precision
SEN: Sensitivity
SPE: Specificity

