## Supplementary for "Novel Large Empirical Study of Deep Transfer Learning for COVID-19 Classification Based on CT and X-Ray Images": Additional File.docx

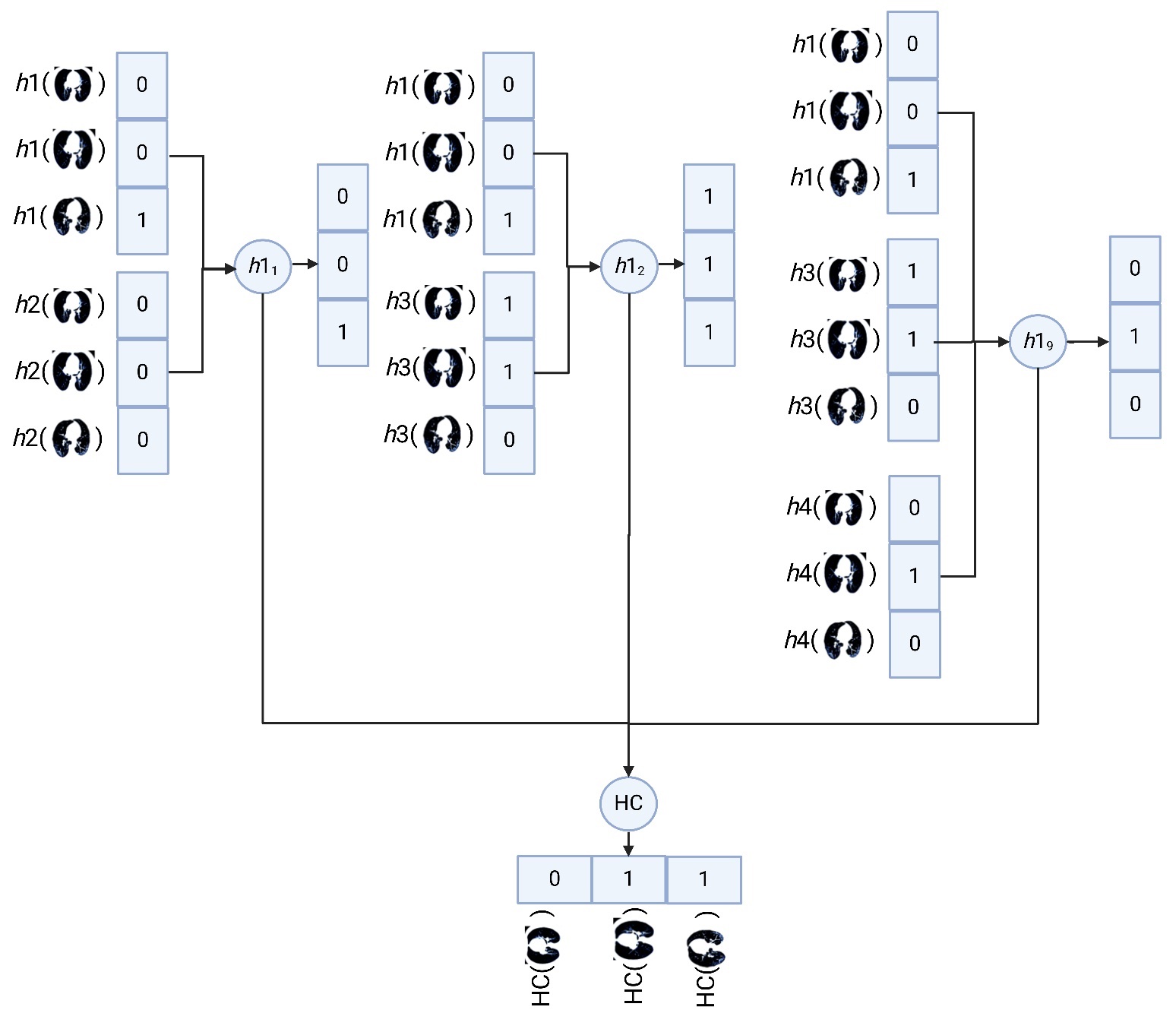

**Figure S1**: Explaining HC model as described in Equations 9-12 performing prediction to three CT images.

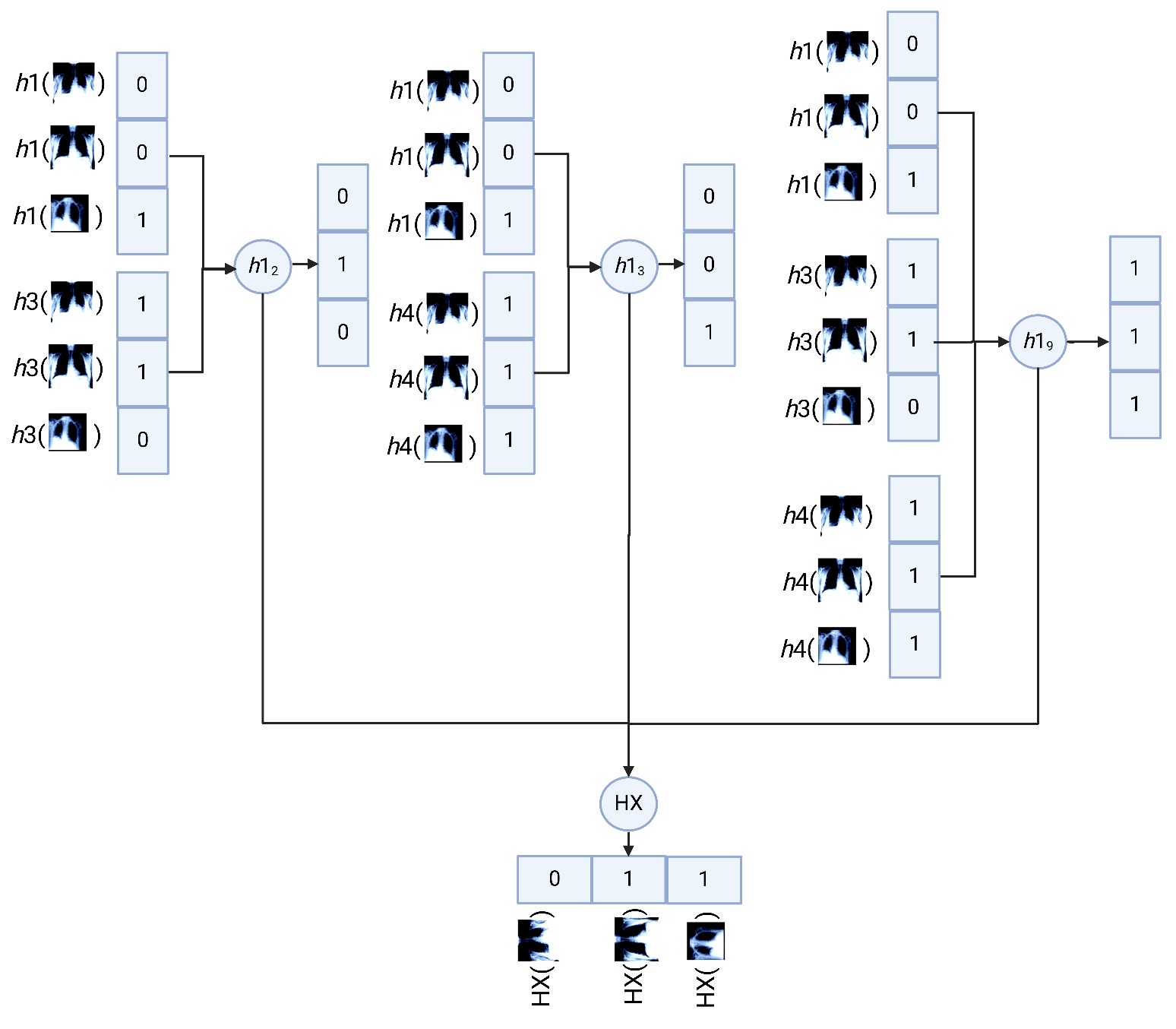

**Figure S2**: Explaining HX model as described in Equations 13-16 performing prediction to three X-ray images.

| **a** | **b** |
| --- | --- |
| 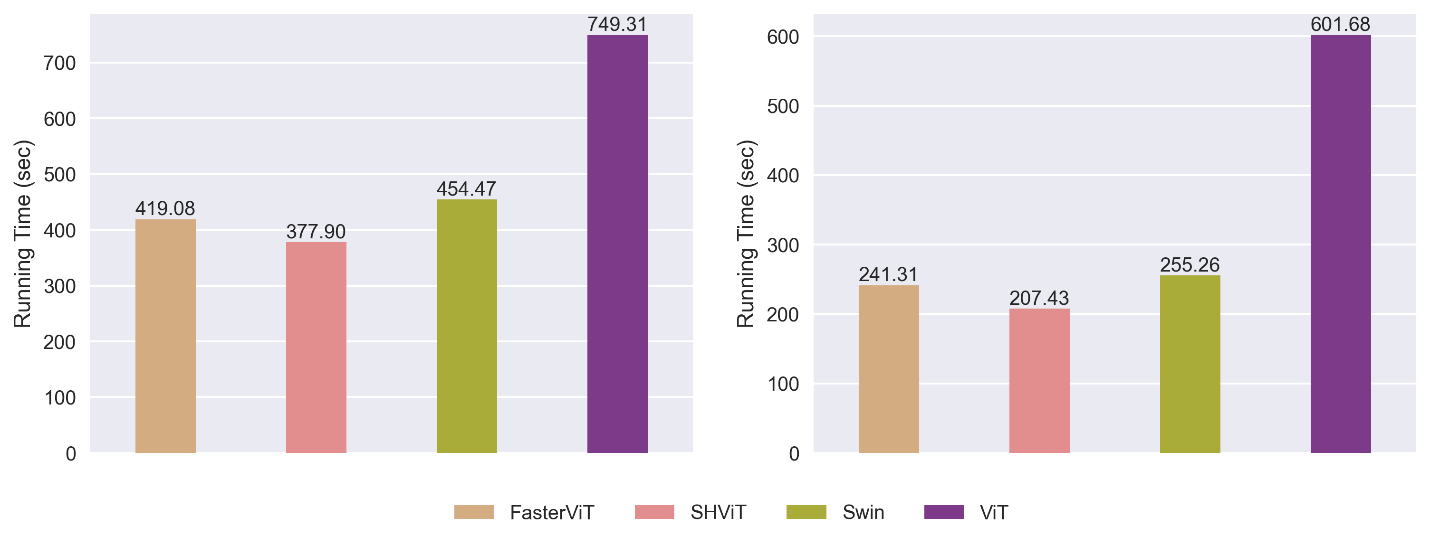  **Figure S3**: Total running time for the transformer-based DTL models during the training phase in five-fold cross-validation pertaining to (a) CT and (b) X-ray datasets. | |

**Table S1**: Summary for layers and parameters pertaining to transformer-based DTL models in this study.

| Model | No. Params | Unfrozen Layers | Transferred | Trainable |
| --- | --- | --- | --- | --- |
| FasterViT | 1,592,129 | 1 | 1,591,616 | 513 |
| SHViT | 6,010,129 | 1 | 6,009,168 | 961 |
| Swin | 27,520,123 | 1 | 27,519,354 | 769 |
| ViT | 85,799,425 | 1 | 85,798,656 | 769 |
