## Supplementary for "Novel Large Empirical Study of Deep Transfer Learning for COVID-19 Classification Based on CT and X-Ray Images": README.txt

ctLGM: folder contains 10000 results pertaining to CT images, described as follows:
Results for 11 DTL models: 4c2_results.csv, 4c3_results.csv, 4c4_results.csv
Results for 2036 DTL models: 11c2_results.csv, 11c3_results.csv, 11c4_results.csv, 11c5_results.csv, 11c6_results.csv,
11c7_results.csv, 11c8_results.csv, 11c9_results.csv, 11c10_results.csv, 11c11_results.csv
Results for 7953 DTL models: 2036c2_7953_results


xrayLGM: folder contains 10000 results pertaining to X-ray images, described as follows:
Results for 11 DTL models: 4c2_results.csv, 4c3_results.csv, 4c4_results.csv
Results for 2036 DTL models: 11c2_results.csv, 11c3_results.csv, 11c4_results.csv, 11c5_results.csv, 11c6_results.csv,
11c7_results.csv, 11c8_results.csv, 11c9_results.csv, 11c10_results.csv, 11c11_results.csv
Results for 7953 DTL models: 2036c2_7953_results

Additional File: a word file containing additional results
